# “When I had concerns about my own patients…I was told to keep quiet”: Moral Injury in the Era of Mandates Among Healthcare Workers in Alberta, Canada

**DOI:** 10.1101/2025.05.27.25328450

**Authors:** Claudia Chaufan, Natalie Hemsing, Rachael Moncrieffe

## Abstract

Between 2021 and early 2022, vaccine mandates in Alberta, Canada, became among the most stringent in the country. This qualitative study explores the lived experiences of healthcare workers (HCWs) following the implementation of Covid-19 vaccine mandates in Alberta’s health sector. It draws on 80 responses to a single open-ended question from a survey of 190 HCWs in the province across vaccination statuses. We performed a manual sentiment analysis, classifying entries as positive, neutral, or negative - depending on their normative orientation towards vaccination mandates - using Weberian ideal types as a conceptual framework.

Most respondents (82.5%) expressed negative sentiments, with close to one fifth (17.5%) offering positive views; no entries were coded as neutral. Themes within negative responses included coercion, ethical conflict, professional exclusion, institutional betrayal, and suppression of dissent. Many vaccinated HCWs described complying under duress, challenging the assumption that uptake signals endorsement. The most salient theme was that of moral injury - defined as the distress caused by acting against one’s conscience, witnessing perceived harm, or remaining silent under institutional pressure. In contrast, positive responses emphasized professional duty and public safety, often rejecting the legitimacy of dissenting perspectives.

Our findings underscore the deeply polarizing nature of vaccination mandates and complicate dominant narratives that equate compliance with consent. Further, in contrast to clinical conceptions rooted in combat or bedside trauma, our analysis situates moral injury in the structural conditions created by public health policies, offering a lens for assessing their implications for the wellbeing of HCWs, quality care, and ethical healthcare practice and policy. We conclude that future public health policies, especially those justified under claims of emergency, must include built-in safeguards for ethical integrity and democratic participation.

---

> Defining general terms is not an abstract exercise but a way of shaping the world metaphysically and structuring the world politically. Wittgenstein’s aphorism, ‘don’t look for the meaning, look for the use,’ is pertinent here. The ethical problem in defining the concept of ‘health’ is to determine what the implications are of the various uses to which a concept of ‘health’ can be put. To carry Wittgenstein a step further, “don’t look for the uses, look for the abuses – [for instance], what can no longer be done in the name of “morality” can now be done in the name of “health”: human beings labeled, incarcerated, and dismissed for their failure to toe the line of “normalcy” and “sanity”.
>
> Daniel Callahan, The WHO Definition of Health (1973)

## Introduction

Prior to the introduction of vaccine mandates, Alberta had one of the lowest Covid-19 vaccination rates - 71% of those who were eligible in Canada - second only to Nunavut (Rieger 2021). During late September 2021, surgeries and other procedures were cancelled due to reported Covid-19 patient loads overwhelming health services, and over 60 infectious disease physicians in Alberta penned an open letter to Premier Jason Kenny, pleading for vaccine mandates because the “healthcare system is truly on the precipice of collapse” (Cecco 2021). In response, Premier Kenney, referring to the unvaccinated, agreed that: “Their choices are now jeopardizing our health care system” (Kaufmann 2021). Thus between 2021 and early 2022, Alberta’s vaccine mandates became among the most stringent in Canada.

According to CBC News (Nov. 13, 2021) and Alberta Health Services, the government implemented a province-wide “restrictions exemption program” that barred unvaccinated individuals aged 12 and up from accessing restaurants, movie theatres, sports venues, and other indoor public settings unless they provided recent negative, “privately paid” Covid tests, or government approved medical exemptions (CBC 2021). Simultaneously, healthcare workers (HCWs) across Alberta were required to be fully vaccinated by October 31 as a condition of employment (CBC 2021), although the deadline was later extended by one month to allow more time for health care settings - particularly long term care settings in rural areas with lower vaccine uptake and ongoing staffing challenges - to prepare for potential labour shortages (Lee 2021). Alberta Health Services’ internal mandate, in place until July 18, 2022, applied to all employees, students, volunteers, and contracted providers. It aimed to “protect patients, healthcare workers and the public” and was, in the words of Alberta Health Services interim Chief Executive Officer Mauro Chies, “the right policy at the time” based on then-available evidence (Alberta Health Services 2022).

As vaccine mandates were slowly dismantled under a new Premier, Danielle Smith, mandate supporters condemned their relaxation by appealing to professional duty and public safety. Referring to the period during which the mandates had been stricter, the Alberta Human Rights Commission stated that mandatory vaccination had been justified, especially during the Delta and Omicron wave, so long as accommodations were made for individuals with disabilities or religious objections, yet clarifying that the Commission “did not deal with personal opinion, political beliefs, the safety or efficacy of vaccines, or other health matters” (Alberta Human Rights Commission 2025). Other supporters warned that reversing mandates could jeopardize longstanding vaccination requirements in healthcare (e.g., for hepatitis B or influenza). For example, a later 2024 article from Canada Health Watch highlighted fears that dismantling mandates in the health sector would lower vaccine uptake beyond Covid, “increasing the risk of preventable disease outbreaks and further destabilizing the province’s healthcare workforce” (Canada Healthwatch 2024). Along similar lines, Lorian Hardcastle, assistant professor in the Faculty of Law and Cumming School of Medicine at the University of Calgary was reported as saying that the amendments could have “more far-reaching impact”, such as preventing the College of Physicians and Surgeons of the province from “disciplining a doctor who speaks out against vaccines”, while Lori Williams, a political science professor at Mount Royal University in Calgary, hypothesized that Premier Smith’s move was “an attempt to tamp down pressure from the vocal and well-organized far-right fringe of her party” (Smith 2024).

Despite this documented initially strong institutional support for mandates, significant dissent emerged throughout this period, both among the public, the healthcare workforce, and different levels of government. For example, tensions were observed between municipal and provincial governments concerning public health mandates and local autonomy, so already in March 2022, the Alberta government passed legislation limiting the ability of municipalities to impose stricter vaccine or mask mandates than the province itself (CBC 2022). As well, the same Alberta Health Services that defended the initial policy, announced in July 2022 that the vaccine mandate was being rescinded, and justified the policy change on grounds of evolving scientific evidence, i.e., that the mandates implemented under the original policy were becoming less effective against Omicron variants, particularly in preventing viral transmission (Alberta Health Services 2022). Premier Kenny then requested that Alberta Health Services identify options for ending the mandate, arguing that “vaccinated health-care workers are pretty much as likely to transmit as unvaccinated and we’re facing workforce challenges, we think that the current policy is not defensible” (Snowdon 2022) while, as noted earlier under Premier Smith, critics counterarguing that the government was appealing to their “voter base” (Cecco 2021; Lee 2022) and that mandates should be maintained in health settings to protect vulnerable patients (Snowdon 2022).

With time, the policy reversal was accompanied by a growing number of legal grievances filed by HCWs. By early 2024, hundreds of them were awarded compensation through union-led arbitration processes. Some were awarded up to $5,000 for being placed on unpaid leave, while others had their testing costs reimbursed - which a limited number of HCWs had incurred as an alternative to vaccination to avoid job loss. Alberta Health Services acknowledged that workers denied accommodations had been placed on leave despite legitimate claims, prompting payouts under the binding recommendations of a mediator (Bruch 2024). Moreover, some legal experts raised concerns about the uneven application of justice. While a small number of HCWs had medical or religious exemptions, the vast majority had been sidelined for asserting personal freedoms or professional judgment, which were not protected under the human rights framework.

These polarized policy shifts - framed initially as necessary to protect the public’s health and later reversed in light of legal, scientific, and political developments - provide the backdrop for this study. Of note, we enter this contested space not to re-litigate the science of vaccines or the legality or politics of mandates - topics that have already been extensively addressed - but to illuminate the lived experience of HCWs in Alberta who were subject to these policies. A growing number of studies, in Canada and beyond, indicate that many HCWs complied under duress, and others experienced exclusion, loss, or moral distress because of institutional enforcement (Chaufan, Hemsing, and Moncrieffe 2024, 2025b, 2025a; McDonald, Parry, and Rhodes 2024). While recent reviews have documented the psychological toll on HCWs of moral harms during Covid-19 (Riedel et al. 2022), these accounts tend to frame them in clinical terms and offer therapeutic remedies. Instead, we approach these harms as an ethical rupture driven by structural conditions, thus requiring not only recognition but importantly institutional reflection, accountability, and change. Our analysis aims to capture the voices of HCWs and map their emotional, professional, and moral landscapes in the context of a rapidly shifting policy terrain.

### Theoretical and Methodological Considerations

We conducted a qualitative reflexive thematic analysis of responses to a single open-ended question (“Are there any other issues that you believe would help us to further understand the impact of vaccine mandates on health workers and patient care? If so, please elaborate”), representing a subset of 80 respondents to a survey of a convenience sample of 190 HCWs in Alberta. Quantitative results from the full survey will be reported separately. Respondents were eligible if they were employed in healthcare or had been employed prior to dismissal due to vaccine mandates. The initial survey was promoted through social media platforms and professional networks of the research team using a snowball sampling approach (Makwana et al. 2023), encouraging participants to share the invitation with other eligible HCWs. All HCWs in Alberta were invited to participate, regardless of vaccination status, profession, age, gender, years of experience, socioeconomic background, or race/ethnicity. Recruitment took place over March and April 2025, with invitations redistributed at seven-day intervals.

Our analysis drew on interpretive sentiment analysis, defined as the study of “opinions, sentiments, evaluations, attitudes and emotions as expressed in natural language text” (Denecke and Reichenpfader 2023). We applied this approach not through computational text mining, as is usually the case, but through manual - typological and thematic - classification. In practice, we categorized entries according to their evaluative stance toward Covid-19 vaccine mandates – explicit or implicit - distinguishing between positive, neutral, and negative sentiments. These categories were conceptualized as Weberian ideal types, heuristic constructs that represent distinct normative orientations (Swedberg 2018) – in our case, toward vaccine policy - and used to assess each response based on its proximity to one of the types, reflecting the respondent’s ethical stance. As well, we adopted a manual, interpretive approach to sentiment coding, in line with literature underscoring the importance of human over machine interpretation in understanding nuanced sentiments (Becker and Sasse 2016; Mattimoe et al. 2021) This framework enabled us to identify broader thematic patterns while attending to heterogeneity within each sentiment category.

Entries were categorized based on both content and tone. Positive entries explicitly supported vaccine mandates, criticized those who resisted them, or affirmed institutional policies with little or no qualification. Negative entries included any form of critique, skepticism, or distress related to the mandates - such as expressions of coercion, regret, ethical tension, professional harm, loss of trust in institutions, or concern for the treatment of unvaccinated colleagues. Responses by vaccinated participants who emphasized informed choice or condemned the marginalization of dissenting HCWs were also categorized as negative, as they reflected disapproval of the mandates despite having complied with it themselves. The neutral category was reserved for responses that lacked a clear evaluative position (Table 1).

**Table 1.** Sentiment Categorization Criteria.

| Sentiment Category | Criteria |
| --- | --- |
| Negative | • Opposes vaccine mandates even if vaccinated<br>• Laments unjust treatment of unvaccinated colleagues<br>• Expresses regret, pressure, coercion, or institutional failure<br>• Advocates for freedom of choice or critiques mandate implementation |
| Positive | • Explicitly supports both vaccination and mandates<br>• Criticizes those who resisted mandates |
| Neutral | • Factual, vague, or lacks evaluative judgment about mandate supporters or dissidents |

After applying this categorization, we thematically analyzed entries (Braun and Clarke 2006) within each sentiment group to identify recurring concepts, experiences, and narrative patterns. Selected entries were included in the results section when appropriate to support our analysis, retaining their original survey identification number. All authors read the full dataset of qualitative entries, and two authors (CC and NH) independently coded the responses and discussed disagreements until consensus was reached.

At this point we should note that while the concept of moral injury was not part of the original design of the study, it emerged inductively during analysis. Defined in the health services literature as the distress that arises from participating in, witnessing, or being unable to prevent actions that violate deeply held ethical or professional values (Litz and Kerig 2019; Shay 2012), moral injury, as a distinct form of moral harm, offers a useful lens through which to interpret many responses in our dataset. Importantly, survey questions were not designed to elicit such specific responses. However, in the course of reading respondents’ testimonies it became apparent that the open-ended format had allowed respondents to voice experiences that may not have been captured through closed-ended prompts. Respondents may have also felt free to share emotionally charged experiences indicating moral injury upon realizing that closed questions in the survey were designed to allow room for dissent without pathologizing it by, for instance, framing it as “vaccine hesitancy” - a largely emotionally driven response in need of a “solution” (Chaufan 2024). The study was approved by the York University Office of Research Ethics (No. 2023-389). The article is presented in accordance with the COREQ reporting checklist.

#### Statement on reflexivity

The research team consisted of three female investigators with over four decades of combined experience in medical sociology and health services research. The lead author is a nonpracticing physician with a Doctorate in Sociology and a Notation in Philosophy, employed as a professor of health policy. The first co-author is a health researcher with a Master of Arts in Anthropology, and the second co-author is completing a Master of Arts in the Health Sciences. Following Malterud (Malterud 2001), these disclosures are offered as a means of enhancing transparency and to ground our commitment to epistemic integrity, rather than for the purpose of personal or ideological positioning. In addition to our academic and professional expertise, our engagement with the topic stems from longstanding concerns about the impact of Covid-19 policies on the healthcare labour force in Canada, as reflected in a substantial body of individual and collaborative research (Chaufan, Hemsing, and Moncrieffe 2024, 2025b, 2025a). Based on our critical policy analysis of the academic literature (Chaufan et al. 2024) this impact has not been adequately addressed in dominant policy narratives.

### Findings

Among the 80 healthcare workers who responded to the open-ended survey question, the overwhelming majority expressed negative sentiments regarding Covid-19 vaccines or mandates. Specifically, slightly over half of entries (41/80; 51.3%) corresponded to unvaccinated respondents and all of them (41/41; 100%) expressed negative sentiments, a small minority (4/80; 5.0%) corresponded to partially vaccinated respondents and all of them (4/4;100) expressed negative sentiments, about a fifth of entries (15/80; 18.8%) were from fully vaccinated respondents with all but one (14/15; 93.3%) expressing negative sentiments, and about one fourth (20/80; 25%) were from respondents boosted once or twice with about one third of these (6/20; 30%), boosted once, expressing positive sentiments, and about two thirds (14/20; 70%), boosted twice or more times, expressing either positive (7/14; 50%) or negative (7/14; 50%) sentiments in equal numbers. While most responses closely aligned with these ideal types, a handful were more ambiguous, yet at close examination, all entries revealed a normative orientation either in support of, or against, the policy of mandated Covid vaccination (Table 2).

**Table 2.**
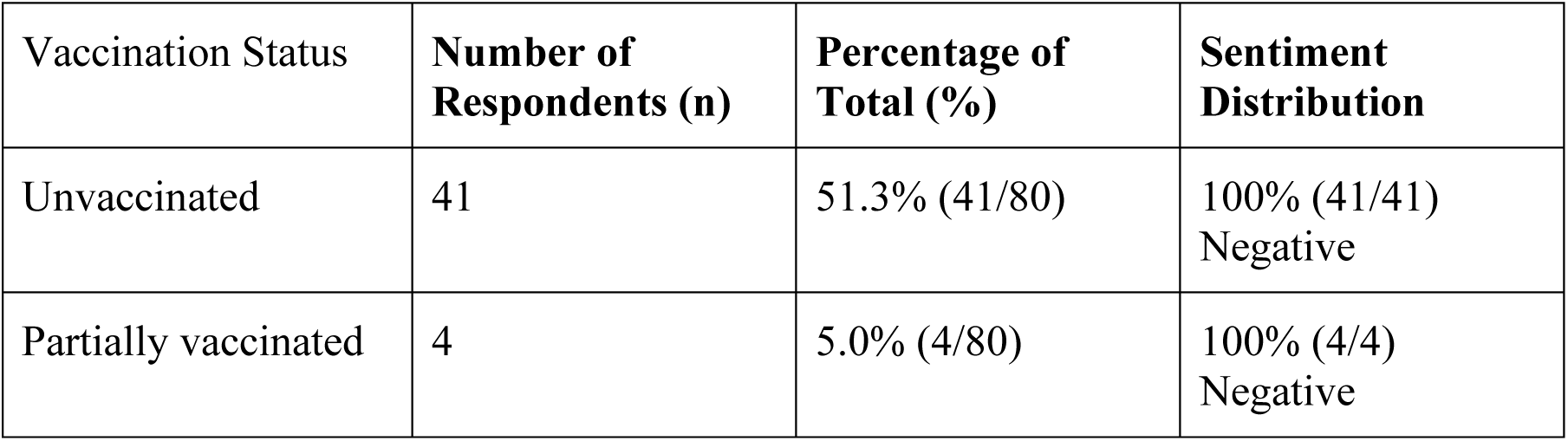
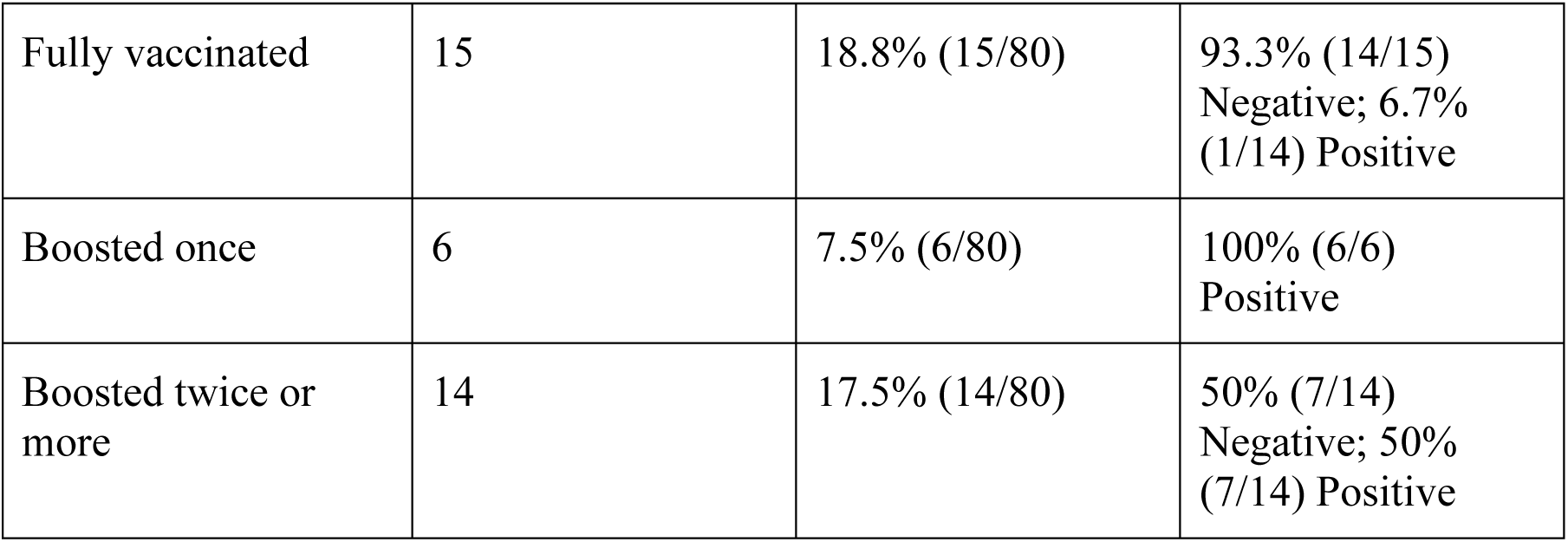
Sentiment Analysis Distribution.

Strikingly, no response was coded as “neutral”, as none fit the description of the category, underscoring the intensity of feeling surrounding HCWs’ experiences of the policy, as well as its polarizing nature, across all groups. We now present and expand on these findings organized by ideal type sentiment.

### Negative Sentiment Toward Mandates

Of the 80 respondents, a substantial majority — 66 entries (82.5%) — expressed negative sentiments toward Covid-19 vaccine mandates. Notably, these responses came not only from unvaccinated and partially vaccinated participants, but also from 14 of 15 fully vaccinated respondents and 7 of the 14 who were boosted twice or more. This distribution highlights that compliance with mandates did not necessarily equate endorsement and that opposition to these policies was widespread across vaccination statuses.

Several themes were identified consistently across these accounts, such as that of the experience of vaccine mandates as *psychologically coercive, emotionally distressing, and ethically troubling*. Many spoke of being forced to choose between their professional identities and deeply held moral or bodily integrity. One fully vaccinated respondent wrote, “I felt completely manipulated and toyed with… I suffer from PTSD and depression from the mandates and subsequent vaccination” (Entry 96). Another recalled complying “at the very last opportunity,” expressing abandonment by both employer and union (Entry 64). These accounts reflect the emotional and psychological toll of navigating high-stakes compliance under threat of exclusion.

Closely linked to these accounts was a profound sense of *violation of bodily autonomy and principles of medical ethics*. Multiple respondents described the mandates as antithetical to the values that healthcare is meant to uphold. An unvaccinated nurse described the policy as “inhumane and unethical… a breach of patient confidentiality, informed consent and bodily autonomy” (Entry 133). Another, who had received the vaccine, criticized what they viewed as ethical inconsistency: “We were hypocritical with our medical ethics… We tell a woman she has a right to abortion… but that same woman should then have no choice in taking a vaccine” (Entry 143). These statements reflect an internalization of professional codes of ethics and a belief that those codes had been violated by institutional policy.

Across the board, participants expressed *a deep sense of institutional betrayal and a collapse of trust* in professional bodies, regulatory colleges, and unions. This theme appeared across most vaccination statuses but was especially pronounced among those who had been dismissed or denied accommodations. A partially vaccinated respondent reflected, “The forcefulness of mandating vaccines removed the choice on own body by losing your job security and has led to many believing that they can no longer trust doctors or government” (Entry 177). An unvaccinated worker described their treatment as “the worst human experience I’ve ever been through” (Entry 90), illustrating the moral weight of what they experienced as punishment, humiliation, and abandonment. These narratives were also marked by *reports of suppression and retaliation*. For example, several participants described being told not to report vaccine-related injuries or being disciplined for raising concerns. One noted simply, “Active discouragement of reporting vaccine side effects” (Entry 180). Others described workplaces where conversation about mandates was tightly controlled, and dissent was punished: “Any discussion around vaccination status or adverse reactions would result in disciplinary action” (Entry 181).

Importantly, not all negative responses came from those who had refused vaccination. Indeed, several vaccinated respondents expressed *solidarity with colleagues who had been marginalized, discriminated against, or dismissed.* One HCW, who had ultimately complied, expressed regret and described ethical tension: “There was a significant lack of support for healthcare professionals that chose not be vaccinated… a rational argument could be made” (Entry 94). These expressions reveal a broader undercurrent of professional and moral concern and complicate assumptions that vaccination equated alignment with the policy. Taken together, these reflections form a coherent and emotionally charged critique of the mandates as unjust, harmful, and at odds with both professional ethics and democratic norms. They also underscore that the boundary between compliance and dissent was not as clear-cut as institutional policies presumed: many respondents complied, but under protest - and with lasting emotional and moral consequences.

#### A rupture in moral orientation: Moral injury in the mandate era

Perhaps the most salient theme within the broader pattern of negative sentiment was the *rupture in moral orientation*, namely, experiences of moral injury, evident in a number of entries from across vaccination statuses. Several participants described being forced to act against their professional or ethical convictions, to remain silent in the face of perceived wrongdoing, or to watch patients or colleagues suffer avoidable harm while feeling powerless to intervene. For example, one fully vaccinated respondent wrote, “I was forced to inject an experimental substance or lose the ability to provide for my family… I wish I had been stronger and refused as well” (Entry 64), reflecting the lasting psychological consequences of perceived ethical compromise. Another respondent, whose comments form the basis of the title of this article, recalled, “I had concerns about my own patients’ negative reactions… But I was expected to keep quiet” (Entry 53), while a third reported, “The guilt rots away at my head” (Entry 90), a phrase that captures the lingering emotional and moral impact of these experiences. Another respondent, reflecting on what they perceived as shifting institutional standards, wrote: “Initially, the vaccines were not approved for pregnant women… then all of a sudden, we were told to vaccinate pregnant women… When I asked for the evidence, I was told there was NONE, and it wasn’t my job to ask questions… I left my job not long after that” (Entry 99). These accounts and others like them point not only to dissent or discomfort but to a more fundamental and enduring rupture: the experience of institutional betrayal and moral dislocation.

### Positive Sentiment Toward Mandates

A small, yet vocal, minority - 14 of 80 respondents (17.5%) - expressed positive sentiments toward Covid-19 vaccine mandates. Notably, all of these responses came from those who were either boosted once or boosted twice or more, with the exception of a single fully vaccinated respondent. These participants framed the mandates not as coercive or ethically troubling, but rather as necessary, professionally responsible, and morally imperative. The most prevalent theme among these responses was a *strong sense of professional duty and alignment with public health ethics*. Participants expressed that being vaccinated - and requiring others to do so - was consistent with the ethical obligations of healthcare work. One respondent emphasized, “I was happy to get vaccinated to protect the vulnerable populations I was working with” (Entry 89), affirming both personal compliance and moral commitment to what they perceived as professionally responsible and socially just. Another elaborated, “As health care workers we are required to show vaccine status… I strongly feel we should support the health of the greater community” (Entry 31). These sentiments reflected not just agreement with mandates and their assumed scientific underpinnings, but the belief that mandates reaffirmed the fundamental norms of healthcare practice and professional ethics.

Another dominant thread was *frustration with unvaccinated colleagues*, who were often portrayed as undermining professional integrity. Some respondents expressed concern or disdain for what they perceived as individualism or irrational fear of Covid vaccines within the workforce. One declared, “Those HCWs who elected not to get immunized… had an impact on me. The lack of accountability as a professional to keep patients safe was deplorable” (Entry 46). Another stated unequivocally, “HCWs who refused to be vaccinated ultimately put their patients at risk which is ethically wrong” (Entry 39). These critiques suggest that for this group, vaccination status had become a litmus test for professional legitimacy and moral integrity.

A striking subset of HCWs supportive of mandates went further, voicing *disapproval of the survey’s openness to dissenting perspectives* - suggesting that this conferred undue legitimacy by implying a moral equivalence with mandate supporters. As one respondent remarked, “These questions have implicit bias that assumes negative experiences” (Entry 59), while another one expressed concern about the very premise of the study: “I actually think this survey is biased towards supporting an assertion that the policies were more harmful than beneficial to workers” (Entry 128). These responses jointly suggest that, among this group, support for mandates was so strongly held that the very act of asking for alternative views was read as an unacceptable, ideological deviation. Overall, the positive sentiment responses were not only aligned with institutional policy but often actively policed the boundaries of acceptable discourse, casting doubt on the legitimacy of critique and reinforcing the view that compliance was scientifically justified and ethically non-negotiable.

### Neutral Sentiment Toward Mandates

None of the 80 responses were categorized as neutral. While some responses contained ambiguous or vague language, every participant ultimately conveyed a clear evaluative stance either in support of, or in opposition to, vaccine mandates - or to the broader institutional and ethical frameworks under which they were imposed. This lack of neutrality underscores the intensely polarizing nature of the mandates and the ethical stakes perceived by respondents.

## Discussion

Our sentiment analysis of 80 respondents within a survey of a convenience sample of 190 healthcare workers in the province of Alberta found that the overwhelming majority (82.5%) expressed negative sentiments, with less than a fifth (17.5%) offering positive responses. No responses were classified as neutral, underscoring the polarizing nature of the policy. Themes within negative responses included coercion, ethical conflict, professional exclusion, institutional betrayal, and suppression of dissent. Many vaccinated respondents described complying under duress, complicating assumptions that vaccine uptake equates endorsement of vaccines, or even mandates, as the literature has often assumed (Chaufan 2023). The most salient theme identified among entries expressing negative sentiment was that of moral injury - the psychological and ethical distress caused by being forced to act against one’s conscience, to witness perceived harm, or to remain silent in the face of perceived injustice. These injuries were not tied to isolated clinical decisions but to broader structural conditions imposed by policy. By contrast, those who supported the mandates often viewed dissenters as unprofessional and interpreted the survey’s openness to criticism of mandated vaccination as bias. Responses within this group appeared to be so strongly held that the very act of exploring alternatives was perceived as inadmissible - outside the boundaries of professional behaviour, civilized discourse, and permissible dissent in a democratic society, such that its suppression became not only acceptable but ethically justifiable.

It should be noted, however, that while this stance could be interpreted as sheer intolerance, especially when expressed toward colleagues who experienced exclusion, job loss, and even deprivation of professional identities, it could instead, or in addition, be read as indicating more complex emotions. For instance, it could be interpreted as an intense moral reaction triggered by the “state of exception” (Agamben 2020) created by the Covid policy response and promoted by a global public health messaging campaign that was not only far-reaching but also overwhelmingly uniform – one that presented mandates as both scientifically necessary and ethically non-negotiable. In such a context, some HCWs likely internalized this framing to the point that rejecting it felt tantamount to abandoning the Hippocratic oath of “primum non nocere” – first do no harm – *by remaining unvaccinated and even failing to recruit others towards embracing vaccination*.

Put another way, what might be read as hostility could also be read as the result of sincere - if institutionally and ideologically shaped – collective moral urgency, as a form of *groupthink*, a term coined in the 1970s to capture a mode of thinking whereby members of cohesive groups tend to align with what they perceive to be the “consensus” within their group, especially in times of geopolitical tension. As Irving Janis noted in the 1970s, this discursive environment - what one might call an ideological battlefield of “good” vs. “evil, or “us’ vs. “them” – constrained the conditions under which dissent could be voiced, or even imagined, during the Cold War era (Janis 1971). Similarly, under the “emergency” projected by the Covid policy response, this “wartime thinking” may have well provided the context for the intense stigmatization against dissenters that ensued: as sociological accounts of stigma and deviance suggest (Link and Phelan 2001), when dissent is framed not merely as error but as danger, the social and professional cost of tolerating it can become prohibitively high.

In sum, these equally plausible, albeit competing interpretations, we believe, jointly shed light on the power of institutional messaging to shape the moral landscape within which HCWs operated in the Covid era - and arguably continues to do so. A case in point is a 2025 document from the Ontario Ministry of Health, which recommends continued vaccination for HCWs to protect vulnerable populations from hospitalization and death (Public Health Agency of Canada 2025). While framed in technocratic terms, this messaging implicitly casts noncompliance as a moral failure. Even in the absence of overt moralizing language, technocratic messaging often conveys deeply normative expectations about compliance and solidarity, particularly when framed as imperative to collective safety – as noted by Daniel Callahan in the epigraph, as a “failure to toe the line” of normative expectations about “normalcy” and “sanity”, in the *medical* and *moral* sense (Callahan 1973).

Our findings also add important nuance to ongoing debates about Covid-19 vaccine mandates in healthcare, in Canada and beyond. First, they problematize the widely held belief that high vaccine uptake indicates acceptance, rather than an inability to refuse a policy whose consequences were too costly for most HCWs to bear. They also position moral injury as a critical lens for understanding the long-term ethical consequences of public health decision-making, especially during emergencies – real or perceived. As mentioned earlier, moral injury has been increasingly acknowledged in the context of frontline trauma care, or even in the context of Covid although typically framed in clinical terms, thus reasonably followed by therapeutic remedies. However, to our knowledge its relevance in the domain of policy compliance, particularly in systems that claim to uphold ethical integrity, remains underexplored. Our study underscores that in the Covid era, moral injury did not arise from bedside dilemmas – at least not only - but rather from the systematic enforcement of policies perceived by many as unethical. Therefore, the accounts in our findings represent not merely testimonies of profound moral dislocation, but most significantly, equally profound indictments of said policies.

At the same time, we acknowledge that the concept of moral injury, while useful, carries certain limitations and even risks. Once adopted into the lexicon of health sciences, there is a risk that moral injury becomes medicalized, i.e., pathologized, thus redefined as a form of psychological or psychiatric dysfunction - hence a form of social control (Conrad 1979) - rather than an expression of systemic ethical rupture (Zola 1975). In such cases, the focus can shift from structural and political responsibility to individual resilience. In our view, the appropriate response to moral injury among HCWs should not be clinical intervention, at least not only, but a structural and political rethinking of public health ethics - one that prioritizes transparency, proportionality, diversity, inclusivity, and respect for fundamental bioethical principles (UNESCO 2005). The harms identified in our analysis are not merely personal but systemic, and the remedy must be similarly collective.

### Limitations and strengths

This study has limitations, many of which are common to qualitative inquiry and others which are specific to the controversial nature of Covid-19 mandates. First, we relied on a non-probability, convenience-based snowball sampling strategy, which limits the generalizability of our findings. However, qualitative research does not aim for statistical generalization but rather transferability - the extent to which findings resonate with or inform other contexts - which we sought to support through thick description and contextual detail (Drisko 2025).

Second, although recruitment was open to HCWs of all vaccination statuses, our sample was skewed toward unvaccinated or vaccine-hesitant participants, many of whom experienced direct punitive consequences from the mandates. As such, their perspectives may not represent the full range of sentiment among HCWs in Alberta. That said, their testimonies offer unique insights into lived experiences that have often been marginalized or excluded from dominant narratives. It also offsets the bias in studies where HCWs compliant with the policy, i.e., vaccinated, are overrepresented. This is the case, for instance, of a systematic review on the attitudes of HCWs towards mandatory vaccination, whose authors reported that their review was potentially biased because included studies referred to mostly vaccinated workers (Politis et al. 2023), a predictable finding given widespread vaccination or termination policies in the industry. In this respect, our study prioritizes ethical representation of harm, rather than proportional representation of majority opinion.

Third, while high vaccination uptake among HCWs is often interpreted as endorsement of vaccine policy, our findings, along with national data, complicates the assumption that compliance indicates consent. For example, a 2023 national survey conducted by Ipsos for the Public Health Agency of Canada found that nearly half of HCWs cited the threat of job loss as a primary motivation for vaccination (Ipsos & Public Health Agency of Canada (PHAC) 2023). These data suggest that coercion played a major role in driving uptake, and that many HCWs vaccinated under duress may not appear in institutional data reporting “uptake”, yet implying “acceptance/endorsement”, as dissenters, even if they are.

Fourth, we note that qualitative research on dissenting perspectives during Covid-19 - particularly those challenging institutional policy - is inherently difficult, given the suppression, stigmatization, and marginalization of such views within mainstream scientific and media discourse. As others have noted (Martin 1999, 2025; Moran 1998; Shir-Raz et al. 2023), experiences that diverge from official narratives are often dismissed or mischaracterized. This context may have contributed to the eagerness of certain HCWs to participate in our study, and to the scarcity of other venues where their perspectives could be shared.

Fifth, it could be argued that this study is of limited utility given its specific Alberta context – a mere 5 million people in a country of about 40 million, a fraction of the world population, even if the Covid event was and remains a global phenomenon. To this objection we would like to present the following observations: across Canadian provinces, the implementation of vaccine mandates for HCWs followed similar timelines and logics, yet their reception and consequences varied. In Ontario, mandates were introduced through Directive #6 and were interpreted by many HCWs as ethically fraught, particularly when framed as “vaccinate or be terminated” policies with minimal avenues for exemption (Chaufan, Hemsing, and Moncrieffe 2024, 2025b). In British Columbia, similar policies extended well beyond the WHO declaration of the end of what the organization had framed as a global health emergency, and were enforced through a rhetorical intensity that painted dissenters as unprofessional or “anti-science” (Chaufan, Hemsing, and Moncrieffe 2025a). In both provinces, many participants reported coercion, professional ostracism, and impacts on patient care that they were not allowed to report.

We argue that what emerges across these jurisdictions is easily transferable to others: a pattern not merely of contested bioethics, but of mandates implemented under what we suggest may be called “democratic coercion” - that is, policy enforcement mechanisms typically associated with authoritarian contexts but made palatable through appeals to public health necessity. While Canada is often framed as a paragon of equity and evidence-informed practice, these cases jointly suggest that, at least in moments of crisis, policies may reflect not deliberative consensus but institutional force. Alberta’s HCWs offer a unique contribution to this evolving picture as their often dissenting voices help us to grasp the emotional and ethical terrain navigated by HCWs more broadly, under extraordinary pressures, including the conflation of compliance with professionalism and ethical behavior.

Finally, we also believe that our study offers several additional important contributions. It provides what is, to our knowledge, the first empirical analysis of moral injury among HCWs resulting from Covid-19 vaccine policy compliance, rather than from clinical trauma or bedside decision-making. By distinguishing between compliance and agreement, the analysis challenges common assumptions about uptake indicating endorsement. Our use of Weberian ideal types to frame sentiment categories, paired with thematic analysis, strengthens both the conceptual and interpretive rigour of the study. The inclusion of vivid, firsthand quotations amplifies the emotional and ethical range of participants’ responses and highlights the polarized nature of the vaccination mandate policy and its impact on the lived experience of the healthcare labour force.

## Conclusion

This study has shed light on the lived experiences of HCWs in Alberta who were subject to Covid-19 vaccine mandates during a period of intense policy enforcement and public health uncertainty - an enforcement that, in many settings, remains ongoing. For example, at the time of this writing some healthcare institutions across Canada continue to require as a condition of employment vaccine doses often administered over three years ago (see for example (Peterborough Regional Health Centre 2025; Ross Memorial Hospital 2025)). This is despite early evidence of widespread community transmission (Bendavid et al. 2021; Majdoubi et al. 2021), benefits largely based on mathematical modelling (Watson et al. 2022) of questionable assumptions (Burch et al. 2024), and the well documented evidence of harms from Covid vaccination (Faksova et al. 2024; Fraiman et al. 2022; Hulscher et al. 2024; Mansanguan et al. 2022; McCullough, Wynn, and Procter 2023; Mead^1^ et al. 2025) – among other considerations. Given that most HCWs, especially those in the frontlines, were exposed from the earliest stages and worked uninterruptedly – often beyond the call of duty - before vaccines existed or were required as condition of employment, these mandates appear not only ethically troubling, but also scientifically incoherent.

By highlighting the diversity of viewpoints across vaccination statuses, our findings also challenge the widespread assumption that compliance implies agreement, as well as the belief that measurable outcomes - such as vaccine uptake - can stand alone as markers of policy success (see for example (Okpani et al. 2024)). As our findings have shown, such outcomes may well conceal the coercive, ethically fraught conditions under which “uptake” was achieved. We therefore conclude that greater attention should be paid to the institutional and political contexts that produce moral harm, and to the voices of those who experience it but are excluded from decision-making processes.

On the positive side, recognizing moral injury as a product of structural and ethical violations - not merely psychological strain - offers a concrete path forward. Future public health policies, especially those justified under claims of emergency, must include built-in safeguards for ethical integrity and democratic participation. This includes meaningful informed consent, proportionate risk communication, and genuine public deliberation. It also requires mechanisms for policy accountability that do not rely solely on appointed experts but include representatives of the public whose lives and livelihoods are most directly affected, and whose interests the appointed experts seemingly represent and serve. Ethical governance cannot remain the domain of expertise - it must be socially grounded and politically accountable. Preventing moral injury in the health sector is not a matter of treating distressed HCWs after the fact, but of crafting policies that minimize ethical coercion. Only then can public health policy advance both its scientific and its ethical commitments.

## Data Availability

Data produced for this present works is protected by a confidentiality agreement with survey respondents

## Acknowledgments

CC thanks the many professional and lay organizations, students, trainees, and friends who have afforded spaces of reflection and debate over the past years, and especially Julian Field, for his editorial feedback and support. NH thanks her family and friends for their encouragement and support, and Dr. Chaufan for her mentorship. RM thanks her friends, family, and Dr. Chaufan for their support and guidance. All authors are grateful to the participants for sharing with us their life experiences and making this study possible.

## Ethical Statement

The authors are accountable for all aspects of the work in ensuring that questions related to the accuracy or integrity of any part of the work are appropriately investigated and resolved. The study was approved by the York University Office of Research Ethics (No. 2023-389).

## Author Contributions (CRediT taxonomy)

Conceptualization: Claudia Chaufan

Methodology: Claudia Chaufan

Writing – Original Draft: Claudia Chaufan

Writing – Review & Editing: Claudia Chaufan, Natalie Hemsing, Rachael Moncrieffe

Investigation: Natalie Hemsing and Claudia Chaufan

Formal Analysis: Claudia Chaufan, Natalie Hemsing & Rachael Moncrieffe

Project Administration: All authors

